# Pilot randomized clinical trial of virtual reality pain management during adult burn dressing changes: lessons learned

**DOI:** 10.1101/2023.03.15.23287329

**Authors:** Megan Armstrong, Rebecca Coffey, John Luna, Henry Xiang

## Abstract

Opioids are the most frequently used pain medications by US burn centers to control severe procedural pain during wound care. Concerns for long-term opioid use have prompted the exploration of non-pharmaceutical interventions, such as virtual reality (VR), for procedural pain management. The primary objective of this pilot study was to evaluate the feasibility and efficacy of VR pain alleviation treatment on reducing adult burn patients’ perceived pain during burn dressing changes. Adult patients aged 18-70 years were recruited from the inpatient unit of a single American Burn Association–verified burn center between May 2019 and February 2020 and randomly assigned to one of three arms. Active VR participants played four VR games; passive VR participants were immersed in the same VR environment without the interaction elements; and a standard of care control group. 71 patients were screened for eligibility and 33 were deemed eligible to approach for informed consent, with 14 agreeing to participate in this study. Of these 14 patients, 4 were randomly assigned to the active VR, 4 to the passive VR, and 6 to the control group. Self-reported overall pain was lowest among participant in the active VR (dressing 1= 41.3, dressing 2= 61.0, and dressing 3= 72.7) and highest among participants in the passive VR (dressing 1= 58.3, dressing 2= 74.5, and dressing 3= 89.0) across all three dressing changes. Self-reported worst pain was lowest among the active VR at the first and last dressing (64.3 and 92.2, respectively), but the control group has the lowest self-reported worst pain at the second dressing (71.3). VR is a useful non-pharmacological tool for pain distraction but designing and implementing clinical research studies face many challenges in real-world medical settings. Lessons from this study have important implications for future VR studies by other researchers.

**Trial Registration:** ClinicalTrials.gov Identifier: NCT04545229

**Author Summary:** In this paper we describe our experience conducting a randomized clinical trial using virtual reality as a pain distraction during inpatient burn care. This pilot study was designed to evaluate feasibility and efficacy of virtual reality as a pain distraction treatment. Three groups intervention groups were compared across multiple burn care procedures. We enrolled 14 patients across 10 months of recruitment. While our sample size was too small to consider significance testing, we did find that the group with active virtual reality participation reported less pain than those in the other two groups. We documented many challenges with using virtual reality during burn dressing changes, including the severity of injuries and the need for high-dose opioids, lack for interest in virtual reality, the unique nature of inpatient wound care, and methods of interacting with a game. COVID-19 also created recruitment restriction for our study. We proposed methods for circumventing these challenges for future researchers when designing virtual reality studies among adult patients.

## Introduction

Burn injuries affect millions of individuals around the world [1]. There were an estimated 204,319 nonfatal burn injuries (about 82 per 100,000) among US adults in 2020 [2]. For moderate to severe burns, daily dressing changes are required, which are very painful and often require high-dose opioids. A 2016 survey by Meyers eta al. of 378 nurses and physicians from 133 American Burn Association Burn Centers reported that for burn dressing changes, oxycodone, morphine, or fentanyl are the most frequently used opioid medications, respectively [3]. This is particularly concerning due to the current opioid epidemic [4]. Meyers et al’s 2016 study found that 80% of respondents reported that procedural burn pain could be more adequately controlled [3].

Researchers have shown that high dose of opioid medication for acute pain management is more likely to increase the risk of long-term opioid use and opioid addictions, for both pediatric and adult populations [5, 6]. Balancing pain management and opioid reduction has been an ongoing challenge in US burn centers. A qualitative study of adult burn patients post-rehabilitation found that adapting to pain was one of the biggest barriers to fully coping emotionally, with failure to cope leading to prolonged depression and anxiety [7].

Increasingly, non-pharmaceutical interventions have been explored for procedural pain management, with virtual reality (VR) being one of these promising interventions [8]. Numerous studies have found that VR significantly decreases procedural pain and anxiety [9-13]. VR is hypothesized to have effective pain management by creating a 3-dimensional distraction that limit the user’s perception of painful stimuli [14]. Overall, VR has very few reported side effects and most participants find the experience enjoyable [9]. VR’s promise with the urgent nature of the opioid epidemic makes studies focused on VR-assisted pain management vital. A recent meta-analysis concluded that while VR intervention could statistically reduce pain intensity among pediatric patients, VR’s effect among adults remains unknown and needs to be investigated [15].

The primary objective of this pilot study was to investigate feasibility and efficacy of VR pain alleviation treatment (VR-PAT) on reducing adult burn patients’ perceived pain during burn dressing changes. We hypothesized that VR-PAT would decrease self-reported pain intensity among inpatient adult burn dressing changes. Findings and lessons from this study have important implications for this strategically chosen research pathway and for future VR studies by other researchers.

## Methods

This three-group pilot randomized clinical trial (RCT) was designed to test feasibility and efficacy of a smartphone VR-PAT in reducing self-reported and observed pain of adult burn injury patients during repeated inpatient dressing changes. Adult patients with burn injuries were recruited between May 2019 and February 2020 from an American Burn Association-verified Burn Center and randomly allocated to one of three treatment arms: active VR, passive VR, or a standard of care. Eligible subjects were 1) 18-70 years of age (inclusive), 2) their first admission for this acute burn injury, 3) require dressing changes, 4) using opioids for dressing changes, and 5) whose burn is ≤4 days from their burn injury. Patients were excluded if they had 1) severe burn(s) on the face/head preventing utilization of VR, 2) cognitive/motor impairment preventing valid administration of study measures, 3) unable to communicate in English, 4) prisoners and patients who were pregnant, and 5) patients admitted to the intensive care unit. The institutional review board of The Ohio State University reviewed and approved this study. Written informed consent was collected from each patient prior to participation in the study. The initial funding of this pilot study met the nonapplicable study definition of the Food and Drug Administration Amendments Act §801 for registration at ClinicalTrials.gov. However, a late voluntary registration was made (ClinicalTrials.gov Identifier: NCT04545229).

This pilot study intended to assess the feasibility of VR-PAT among hospitalized adult burn patients, so formal power and sample size calculations were not conducted. Instead, the intention was to generate preliminary data to estimate the effect size of VR and variances of the outcome measures that could be used to calculate the sample size and power of a future larger-scale randomized trial. Based on our previous VR study among pediatric patients and the availability of adult burn patients at the adult burn center, the planned enrollment was 60 patients equally randomized into three groups (active VR, passive VR, and standard of care). A total of 71 patients were screened for eligibility and 33 were deemed eligible to approach for informed consent, with 14 agreeing to participate in this study (**Figure 1)**. Of these 14 patients, 4 were randomly assigned to active VR, 4 to passive VR, and 6 to the control group. Patient recruitment was stopped in March of 2020 because of the hospital restrictions in response to COVID-19 and the future uncertainty promoted the funding agency to withdraw the funding soon after.

**Figure 1.**
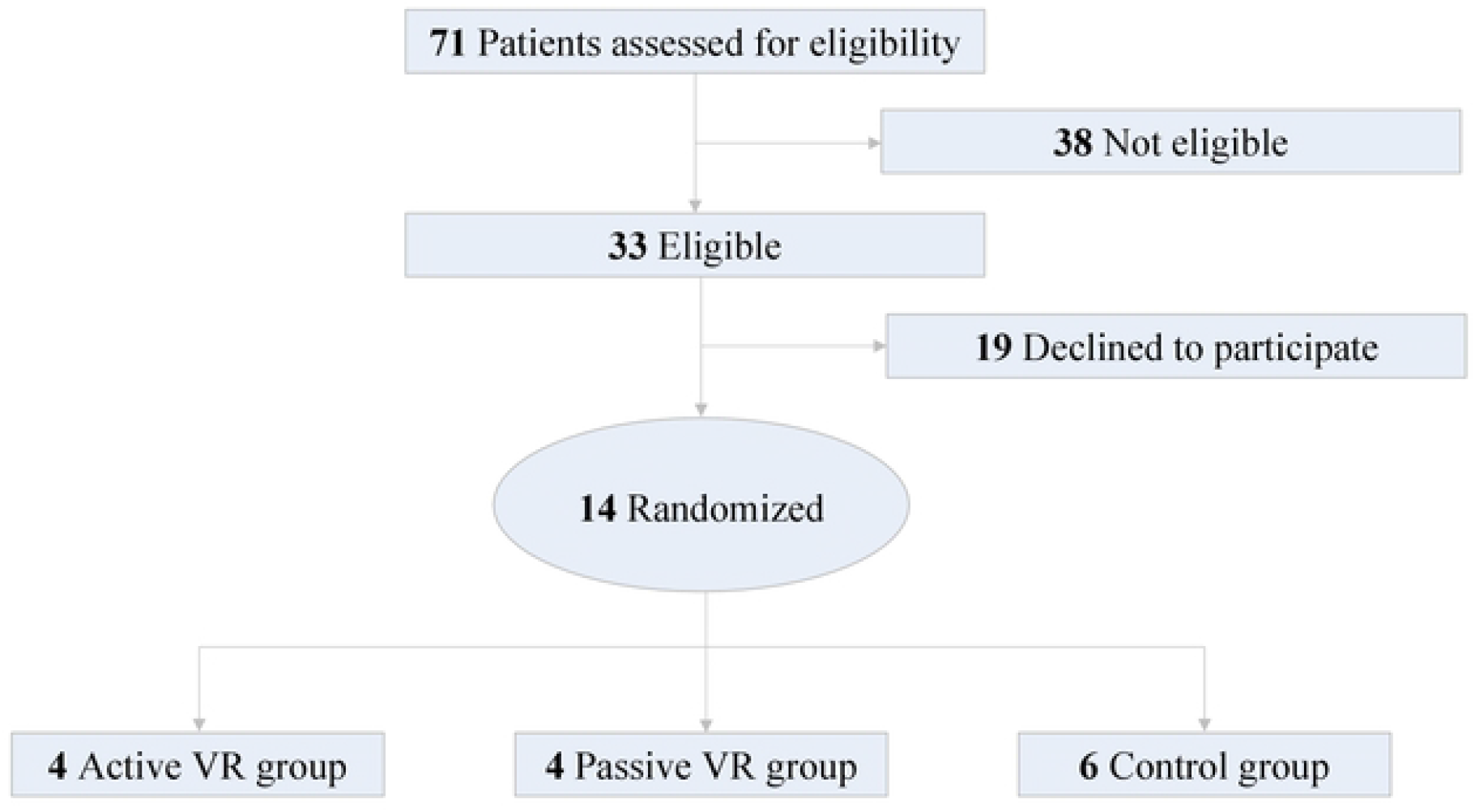
Screening and Recruitment Diagram

### Study Procedures

Participants were identified by the physicians overseeing their burn care and approached by a researcher trained for this study, prior to their next inpatient dressing change. Following informed consent, participants were randomized to one of the three intervention groups using a 1:1:1 allocation ratio, which was hosted on a Research Electronic Data Capture (REDCap) [16, 17] site. The researcher recruiting the subject was blinded to the randomization sequence. The clinicians filled out a “Participant Information Sheet” following recruitment, which contained demographic and injury variables. Clinicians also documented the subject’s daily opioid use using morphine milligram equivalents (MME). Immediately prior to the dressing change, the research associate completed the “Pre-Intervention Assessment” with the subject and set-up the assigned distraction tool. During the procedure, the research associate completed the “Observed VR Experience” and monitored the use of the device. Immediately following the dressing change, the subject answered the “Post-Intervention Assessment”, and the “VR Experience” survey and nurses completed the “Nurse Perceived Utility” survey. This process was completed once per day for up to three inpatient dressing changes. A follow-up was scheduled 2-6 weeks after discharge where clinicians asked about opioid use and how many pills were consumed in the past 24 hours. This information was then used to calculate 24-hour MMEs.

#### Participant Information Sheet (clinician documented–all subjects)

Using the electronic medical record, clinicians documented demographics (age, sex, and race/ethnicity), injury information (burn date, % total body surface area (TBSA), burn severity (%TBSA that was partial and full thickness), burn location and any illicit drug use prior to injury.

#### Pre-Intervention Assessment (self-reported–all subjects)

Participants self-reported their expectations for the dressing change (How much would you like something fun to do during the dressing change? and How much do you think having something to do during the dressing change would be helpful?) on a visual analog scale (VAS; range 0-100, with higher scores indicating higher expectations). Subjects assessed their state anxiety using the 20-question Spielberger State-Trait Anxiety Inventory (STAI) [18, 19], rated on a 4-point Likert scale (higher scores indicated greater anxiety).

#### Observed VR Experience (researcher documented–all subjects)

Researchers observed the dressing change and documented the total time of the dressing change and VR use, number of times the participant voluntarily interrupted the VR, whether the participant was fully engaged with the VR, and whether they appeared distracted.

#### Post-Intervention Assessment (self-reported–all subjects)

Participants self-rated overall pain, worst pain, time spent thinking about pain, and how much the wound bothered them on a VAS (higher score indicate more pain) and perceived length of dressing (in minutes). Those using the VR were also asked to report any symptoms using the Simulator Sickness Questionnaire (SSQ; lower scores indicate less sickness) [20].

#### VR Experience (self-reported–VR groups)

Participants were asked if they were happy with the VR, whether it could be made better, and if they would want to use it again in the future. They were also asked to report how much fun they had and how engaging they thought it was on a VAS (0-100, higher score indicated more fun and engagement).

#### Nurse Perceived Utility (nurse reported–VR groups)

For patients using the VR, the nurse performing the dressing change reported whether the VR-PAT was helpful and easy to use on a VAS (0-100, higher score was better).

#### Daily Opioid Use (clinician documented–all subjects)

Clinicians documented the MMEs consumed by the participant on each day of participation.

#### Follow-up Assessment (self-reported – all subjects)

A clinician called participants 2-6 weeks following discharge as part of clinical care and asked whether they were still using opioid medications to control pain and calculated the 24-hour MMEs.

### Interventions

VR-PAT was administered using a lightweight, low-cost VR headset paired with an Apple iPhone XS and Bluetooth gaming controller (**sFigure 1**). VR-PAT is a stand-alone game that was developed by Nationwide Children’s Hospital, an affiliated hospital of The Ohio State University.

#### Active VR-PAT

Active VR participants played a series of four VR games specifically designed for this study. These games were broken down into two categories: projectile games (Town and Cave) and rhythm games (City and Forest) (**sFigure 2**). The games played in the order of City, Cave, Forest, and Town for all participants. Participants interacted with the game by tilting their head and pressing a button on the gaming controller.

**Figure 2A&B.**
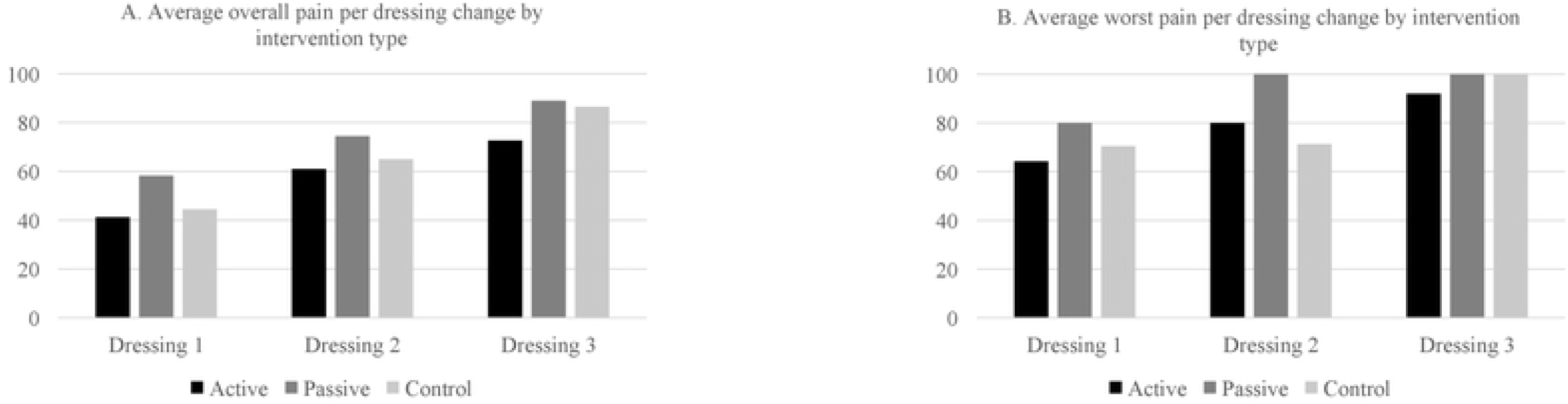
Self-reported overall and worst pain by dressing change and intervention type

##### Projectile games

During the Town game, the user drove around the town in a car and delivered pizzas to the hungry townsfolk by launching the pizzas. During the Cave game, the user drove down a river within a cave full of rats riding in hot air balloons. The user launched rubber balls at the pizza boxes hanging from the hot air balloons to knock them away from the rat bandits.

##### Rhythm games

The City game had users sitting in a car and timing their car horn to have a bird relieve itself on a passerby under the tree. The Forest game had users driving a car through a forest and timing their windshield wiper swipes to prevent the birds from hitting the windshield. *Passive VR-PAT* Passive VR participants went on an automated virtual tour of the same virtual environments as the active VR but were not presented with any of the interactive elements of the games.

#### Control

The standard care participants received routinely used distraction tools (i.e., music and/or talking) provided in the clinical setting.

### Statistical analysis

Demographic and burn characteristics were described using frequencies and percentages for the categorical variables and median and range for continuous variables. Mean was calculated for the primary outcome of reported pain (overall pain, worst pain) across dressing changes and intervention type. VR usage was evaluated by the median length of the dressing time (in minutes) and time using the VR (in minutes). Opioid medication was converted to MME for each medication per 24-hours of admission and added together to get a total amount of opioids.

## Results

Of the 14 participants in this study, they were a median 38.4 years of age, mostly male (71.4%), and White (85.7%) (**Table 1**). Burn injuries varied from 1.0 to 17.8% TBSA (median 8.4%) and the largest proportion of the burns were partial thickness (median TBSA=8.4%). The largest full thickness burn was 3.0% TBSA.

**Table 1.** Demographics and burn characteristics of study participants

| Characteristic | N (%) |
| --- | --- |
| Total | 14 (100%) |
| Age at enrollment, median (range) | 38 (19-70) |
| Sex |  |
| Male | 10 (71.4%) |
| Female | 4 (28.6%) |
| Race |  |
| White | 12 (85.7%) |
| Black | 2 (14.3%) |
| Illicit drug use prior to injury |  |
| Yes | 3 (21.4%) |
| No | 11 (78.6%) |
| VR group |  |
| Active | 4 (28.6%) |
| Passive | 4 (28.6%) |
| Control | 6 (42.9%) |
| TBSA %, median (range) | 8.4 (1.0-17.8) |
| Burn depth, median (range)* |  |
| Partial thickness % | 8.4 (0.5-17.8) |
| Full thickness % | 0.0 (0.0-3.0) |

Abbreviations: VR=Virtual Reality, TBSA=Total Body Surface Area
\*Burn depth was reported as the TBSA% of partial and full thickness for each burn injury

Average self-reported overall pain was lowest among participant in the active VR group [dressing 1= 41.3, dressing 2= 61.0, and dressing 3= 72.7 (mean VAS score)] and highest among participants in the passive VR group (dressing 1= 58.3, dressing 2= 74.5, and dressing 3= 89.0) across all three dressing changes (**Figure 2)**. Average self-reported worst pain was lowest among the active VR group at the first and last dressing (64.3 and 92.2, respectively), but the control group had the lowest self-reported worst pain at the second dressing (71.3). Again, the passive group reported the highest worst pain across the first two dressings (80.0 and 100.0, respectively) and tied for the highest worst pain with the control group at the last dressing (100.0).

Dressing 1 and 2 was longer for the control group (median 59 and 60 minutes, respectively) than the active and passive groups (median 49.5 and 44 minutes and 52.5 and 56.5 minutes, respectively) (**Table 2**). Conversely, the control group used the least opioids for the first two dressings (56.25 and 19.0 MME, respectively) and the passive group used the most opioids for the first two dressings (106.25 and 91.5 MME, respectively). In comparison, the control group had the shortest dressing time for dressing 3 (median 49 minutes) compared to the active and control groups (median 51 and 64 minutes, respectively).

**Table 2.** Virtual Reality (VR) usage and morphine milligram equivalents (MMEs) by participant and dressing numbers

| Subject ID | Dressing #1 |  |  |  | Dressing #2 |  |  |  | Dressing #3 |  |  |  |
| --- | --- | --- | --- | --- | --- | --- | --- | --- | --- | --- | --- | --- |
|  | Days since injury | Length of dressing (minutes) | Time using VR (minutes) | MME | Days since injury | Length of dressing (minutes) | Time using VR (minutes) | MME | Days since injury | Length of dressing (minutes) | Time using VR (minutes) | MME |
| **Active** |  |  |  |  |  |  |  |  |  |  |  |  |
| #3 | 1 | 50 | ** | 90 | 2 | 38 | 15 | 7.5 | - | - | - | - |
| #5 | 1 | 71 | 9 | 98.5 | 2 | 60 | 29 | 97 | 3 | 88 | 26 | 114 |
| #10 | 0 | 10 | 11 | 18 | 1 | 33 | 22 | 22.5 | 2 | 18 | ** | 15 |
| #13 | 0 | 49 | 37 | 83 | 1 | 50 | 22 | 61.2 | 2 | 51 | 25 | 82.5 |
| Median |  | 49.5 | 11 | 86.5 |  | 44 | 22 | 41.85 |  | 51 | 25.5 | 82.5 |
| **Passive** |  |  |  |  |  |  |  |  |  |  |  |  |
| #4 | 1 | 34 | 32 | 120 | - | - | - | - | - | - | - | - |
| #6 | 1 | 77 | 35 | 97 | 2 | 85 | 19 | 78.5 | 3 | 104 | 46 | 78.5 |
| #9 | 1 | 64 | 29 | 115.5 | - | - | - | - | - | - | - | - |
| #14 | 0 | 41 | 15 | 61 | 1 | 28 | ** | 104.5 | 2 | 24 | 9 | 104.5 |
| Median |  | 52.5 | 30.5 | 106.25 |  | 56.5 | 19 | 91.5 |  | 64 | 27.5 | 91.5 |
| **Control** |  |  |  |  |  |  |  |  |  |  |  |  |
| #1 | 1 | 85 | N/A | 86 | - | - | - | - | - | - | - | - |
| #2 | 1 | * | N/A | 65.5 | - | - | - | - | - | - | - | - |
| #7 | 1 | 69 | N/A | 47 | 2 | 81 | N/A | 19 | 3 | 40 | N/A | 32 |
| #8 | 1 | 41 | N/A | 15.5 | - | - | - | - | - | - | - | - |
| #11 | 1 | 51 | N/A | 35 | 2 | 60 | N/A | 7.5 | - | - | - | - |
| #12 | 1 | 59 | N/A | 162.5 | 2 | 45 | N/A | 88.5 | 3 | 58 | N/A | 96 |
| Median |  | 59 | N/A | 56.25 |  | 60 | N/A | 19 |  | 49 | N/A | 64 |

\*Missing
\*\*VR not used for this dressing
N/A –VR not used in control group
MME = Milligram Morphine Equivalents

## Discussion

This pilot RCT study showed some promising results of using VR pain management during adult burn dressing changes. While this study had a small sample size caused by the patient recruitment interruption due to COVID-19, the preliminary results suggest that patients in the active VR group experienced less pain than those in the passive VR and control groups. More importantly, this pilot study had many lessons that could be important to other researchers who plan to design future adult VR pain management studies.

Our preliminary results follow the same trend as our previous VR study during outpatient pediatric burn dressing changes, where participants in the active VR group reported significantly lower overall pain and worst pain [12]. These results are also consistent with a recent systematic review and meta-analysis by Norouzkhani et al. where they reported that the VR-based intervention significantly reduced pain compared to those in the control group [15]. It should be noted that the average age of our RCT was older than that reported in either of these two studies.

All subjects in this study did not require 3 dressing changes and three participants in the VR groups chose not to use their VR for every dressing change. Additionally, as shown in the difference between the time in the VR and the time of the dressing, participants did not wear their VR for the entire dressing procedure. Some participants became bored with the games (active) or video (passive) and did not want to participate for the full dressing. Additionally, some patients began the dressing change with a shower to loosen the dressing, which we found not to be conducive with the VR-PAT process.

### Lessons Learned for Future Studies

First, adults were less interested in participating in VR research than pediatric patients. Some adult patients felt distraction was not necessary or that they did not want to bother with learning how to play the game. Several adults also did not want to be in the virtual VR environment because of motion sickness. Over 53% of the patients screened for eligibility were not eligible to participate, with most of these patients being over the age limit, having facial burns, presented too many days out from the initial injury, and being admitted to the intensive care unit.

The second lesson was that the inpatient adult burn injuries were more severe than encountered in our pediatric outpatient study. These inpatient dressings were also longer than those in our pediatric pilot study, resulting in some participants feeling that the games were repetitive and choosing not to wear the VR for the entirety of the dressing or requesting to not to participate in the VR groups in the next dressing. Suggestions were for more levels or increased gamification to be entertaining for a longer period. Additionally, because of the increased burn severity, the dressing change was more painful, requiring more pre-procedural pain medications and rescue opioid medications. This increased pain broke through the distraction, leading us to hypothesize there is a pain threshold for where VR distraction can be beneficial. This is an important consideration for future VR studies. Future research needs to determine where this pain threshold is and whether a more difficult game can circumvent this issue. Our game was designed to be easy for participants of all ages and skill levels to avoid any feelings of frustration or discouragement. On the other hand, the feedback of increasing the difficulty or adding more levels is good advice for future research.

The third lesson was the unique circumstance that many inpatient dressing changes began with a shower to loosen the previous dressing. We did ask the first few participants to use the VR-PAT during this part of the procedure as the phone and headset were water resistant.

However, participants began experiencing nausea and headaches during this part of the procedure, which medical professionals felt could have been exacerbated by the pain of removing the dressing and the heat/steam of the shower. Upon this realization, the study procedures were changed to only using the VR after the shower portion of the procedure, which did help to reduce these feelings. Additionally, a portion of the participants required additional rescue medications during the procedure, which had their own host of side effects that were similar to simulator sickness symptoms. It was difficult to parse out who was reporting simulator sickness symptoms due to the medication or the VR-PAT, as even participants not in the VR groups reported symptoms. There was less reliability in reporting of all questionnaires among those who required additional medication, with some questionnaires being left blank.

The fourth challenge was the difficulty of using a controller for the game play. As the game play only required the pressing of one button and no movement (i.e., the controller could rest on the bed with only one finger being used), we did not expect it to disturb the dressing change.

However, due to the location of some injuries, participants needed to roll to one side or switch hands when both were burned, and this caused game interruptions. Future studies should consider a hands-free VR system to minimize hand involvement and interference with the dressing procedure.

The final challenge was the COVID-19 pandemic. Study recruitment was paused in March 2020 in compliance with hospital policies. Our funding period ended before recruitment could recommence, thus further limiting our study participants.

## Conclusions

VR is a useful non-pharmacological tool for pain distraction but designing and implementing clinical research studies can face many challenges in the real-world medical settings. Future studies should consider these challenges when working with adult burn patients and work closely with clinic staff and patients to pilot test study protocols before launching a full-scale VR intervention study.

## Data Availability

Even upon de-identifying this human subjects data, it is possible that subjects could be identified due to the small sample size and the one recruitment location. Data cannot be publicly shared, but data sets can be made available upon request

## Acknowledgements

We appreciate Mr. Jonathan Lun for his assistance with the literature review for this project as a summer intern.

**sFigure 1**. Demo of research intern using virtual reality game

**sFigure 2**. Virtual reality game photos and explanations of game concept

